# Aerosol-spread during chest compressions in a cadaver model

**DOI:** 10.1101/2020.03.31.20049197

**Authors:** Matthias Ott, Alfio Milazzo, Stefan Liebau, Christina Jaki, Tobias Schilling, Alexander Krohn, Johannes Heymer

## Abstract

**Objective:** To evaluate aerosol-spread in cardiopulmonary resuscitation (CPR) using different methods of airway management. Knowledge about aerosol-spread is vital during the SARS-CoV-2-Pandemic.

**Methods:** To evaluate feasibility we nebulized ultraviolet sensitive detergents into the artificial airway of a resuscitation dummy and performed CPR. The spread of the visualized aerosol was documented by a camera. In a second approach we applied nebulized detergents into human cadavers by an endotracheal tube and detected aerosol-spread during chest compressions the same way. We did recordings with undergoing compression-only-CPR, with a surgical mask and with an inserted laryngeal tube with and without a connected airway filter.

**Results:** Most aerosol-spread at the direction of the provider was visualized during compression-only-CPR. The use of a surgical mask deflected the spread. Inserting a laryngeal tube connected to an airway filter lead to a remarkable reduction of aerosol-spread.

**Conclusion:** The early insertion of a laryngeal tube connected to an airway filter before starting chest compression may be good for two things – the treatment of hypoxemia as the likeliest cause of cardiac arrest and for staff protection during CPR.

## Introduction

The Coronavirus Disease 2019 (COVID-19) spread all over the world and became pandemic ^1^. There is a high risk of infection with severe acute respiratory syndrome coronavirus 2 (SARS-CoV-2) during patient care for medical professionals, particularly in aerosol-generating procedures such as endotracheal intubation and cardiopulmonary resuscitation (CPR)^1–3^. Data from China, where SARS-CoV-2 emerged, the spectrum of disease (N = 44415) was severe in 14% (6168 cases) and critical in 5% (2087 cases) ^4^. In Italy, 9% (2026 cases) of all COVID-19 cases reported in March (22512 cases) were health care workers^5^. Adequate protection especially during aerosol-generating procedures is of utmost importance. Currently there is little data regarding safety precautions during cardiopulmonary resuscitation. There are different recommendations for staff protection during CPR, e.g. using a face mask or an oxygen mask on the patients face ^6^. A rapidly insertion of a supraglottic airway management device may also protect the staff from spreading aerosol of the virus. The purpose of this study was to visualize aerosol-spread during CPR with different strategies of staff protection and airway management.

## Methods

### CPR Simulation Model

To see whether visualizing of aerosol during chest compressions is feasible, we created an ex situ simulation model using a modified resuscitation dummy. Therefore, a difficult airway trainer was connected to two similar bag valve masks for children to simulate the lung laying in a chest compression trainer. At the end of the host system a nebulizer for inhalation was connected. To visualize aerosol-spread we nebulized different disinfection detergents detectable by ultraviolet light (Ecolab magic blue, Ecolab Deutschland GmbH, Monheim am Rhein, Germany; Bode visirub conc., Bode Chemie GmbH, Hamburg, Germany, Schuelke S&M Optik, Schuelke & Mayr GmbH, Wien, Oesterreich). While applying an air flow of 8 l/min to the nebulizer, chest compression on the resuscitation dummy was performed. The data from this feasibility study are currently under review.

### Cadaver model

In a second approach we visualized aerosol-spread during CPR in a human cadaver model by a cooperation with the Institute of Clinical Anatomy and Cell Analysis, Eberhard Karls University Tuebingen. The human cadavers have been fixed with an ethanol-based technique. To fill the airway and lungs with ultraviolet sensitive detergent, we used a self-inflating bag connected to a nebulizer set (DAR Nebulizer Set, Covidien Ireland Limited, Tullamore, Ireland).

### Data collection and study protocol

We documented the spread of the luminescent aerosol by taking photos in a dark room with ultraviolet lamps. The aerosol, simulated by nebulized ultraviolet sensitive detergents, spreading during ongoing chest compressions was photo documented in different scenarios. At first, aerosol-spread was detected during compression-only-CPR. In a second approach, we attached a face mask on the face. Afterwards we inserted a laryngeal tube (LTS-D, VBM Medizintechnik GmbH, Sulz a. N., Germany) with and without an airway filter (DAR Adult-Pediatric Electrostatic Filter HME, Covidien Ireland Limited, Tullamore, Ireland). Since three cadavers were available for our study, we could only demonstrate limited techniques on the cadaver model due to the rapid deterioration of the mechanical properties of the lung during CPR. We evaluated the extend and direction of the aerosol-spread by visual estimation based on the acquired pictures.

## Results

In our study most spread particularly into the direction of the provider of chest compression provider was recognized during compression-only-CPR without any airway device. Use of a face mask led to deflection of the aerosol. Using a laryngeal tube without an airway filter led to direct spread in the direction of the tube. When a laryngeal tube with airway filter was used, almost no aerosol-spread was visible.

Figure 1 shows visualized aerosol-spread during CPR in different settings.

**Figure 1.**
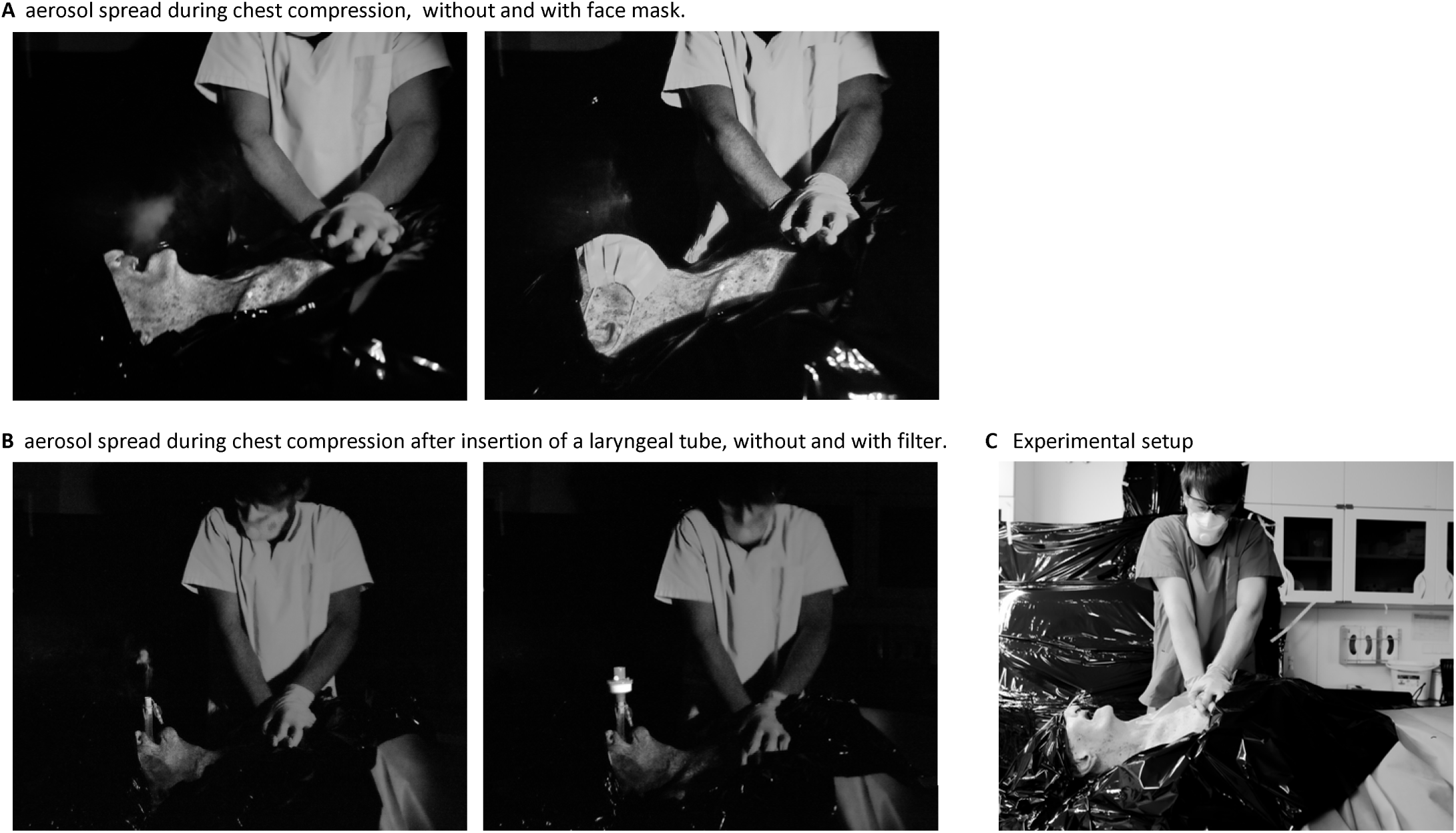
Spread of aerosol in CPR. **Panel A** shows the aerosol spread under chest compression and its reduction when a face mask is applied to a human cadaver. **Panel B** shows the aerosol spread after insertion of a laryngeal tube, without and with airway filter. **Panel C** shows the experimental setup.

## Discussion

COVID-19 is a highly contagious disease with a large number of patients require intensive care medicine^2,4,7^. Critical care is an integral component in patient treatment but contains aerosol-spreading procedures which potentially lead to infection of medical professionals^7^. At this juncture there is little data or recommendation for CPR in times of COVID-19. The Resuscitation Council UK recommends using a face mask for oxygen therapy to minimize aerosol-spread during CPR^6^. In our simulation, applying a surgical face mask leads to deflection of simulated aerosol but seems not sufficient. Our findings suggest that the early insertion of a laryngeal tube connected to an airway filter during CPR reduces the aerosol-spread. Since a lot of inpatient cases develop an acute respiratory distress syndrome (ARDS), early airway management and oxygenation are the mainstay of the treatment for acute deterioration or cardiac arrest^8^. Endotracheal intubation in COVID-19 patients is a high risk procedure and should be performed by specialists with maximum expertise to reduce the risk of infection^1,3,6,9^. Furthermore, bag mask ventilation also seems to have a high risk for infection and should be avoided^3^. The early insertion of a laryngeal tube by any advanced life support provider particularly for staff protection may be an alternative in the setting of cardiac arrest. Performance of airway management - even before starting chest compression – is standing in contrast with actual guidelines of the European Resuscitation Council^10^. Early airway management in cardiac arrest of COVID-19 seems to be reasonable since literature names respiratory failure the main cause of death in these patients^11^. In our simulation model, we used continuous air flow on the nebulizer to simulate aerosol-generation. To create realistic conditions at the cadaver model, we filled the residual volume with aerosol and did not apply any continuous flow. Both components of our experimental setup showed very similar results, making us confident about the validity of the CPR simulation model. Nevertheless, we do not know how close we can get to realistic conditions using an aerosol-spreading model like this because of lack of information regarding this issue. Further investigations as for instance measurements of environmental virus particles may generate more precise answers about contagiosity during CPR.

## Conclusion

CPR and airway management are high risk procedures regarding potential infection with SARS-CoV-2. After visualizing aerosol-spread by ultraviolet sensitive detergents during CPR on a cadaver model, we state that the early insertion of a laryngeal tube connected to an airway filter before starting chest compressions may be a good method for both – treating potential hypoxemia and protect the advanced life support providers. The utmost importance of personal protection equipment stays beyond any debate.

## Data Availability

All original data is available with the author, no online database has been used.

## Conflict of interest

none

## Notes

### Competing Interest Statement

The authors have declared no competing interest.

### Funding Statement

No Funding has been received

## References

1. Alhazzani W, Møller MH, Arabi YM, et al. Surviving Sepsis Campaign: Guidelines on the Management of Critically Ill Adults with Coronavirus Disease 2019 (COVID-19). Crit Care Med. 2020;Online Fir. doi:10.1097/CCM.0000000000004363

2. Zuo M-Z, Huang Y-G, Ma W-H, et al. Expert Recommendations for Tracheal Intubation in Critically ill Patients with Noval Coronavirus Disease 2019. Chinese Med Sci J = Chung-kuo i hsueh k’o hsueh tsa chih. 2020. doi:10.24920/003724

3. Chun-Hei Cheung J, Tin Ho L, Vincent Cheng J, Yin Kwan Cham E, Ngai Lam K. Staff safety during emergency airway management for COVID-19 in Hong Kong. Lancet Respir. 2020. doi:10.1016/S2213-2600(20)30084-9

4. Wu Z, McGoogan JM. Characteristics of and Important Lessons From the Coronavirus Disease 2019 (COVID-19) Outbreak in China: Summary of a Report of 72C314 Cases From the Chinese Center for Disease Control and Prevention. JAMA. February 2020. doi:10.1001/jama.2020.2648

5. Livingston E, Bucher K. Coronavirus Disease 2019 (COVID-19) in Italy. Jama. 2020. doi:10.1001/jama.2020.4344

6. Resuscitation Council (UK). Guidance for the resuscitation of COVID-19 patients in hospital. 2020-03-19. https://www.resus.org.uk/_resources/assets/attachment/full/0/36100.pdf. Published 2020. Accessed March 19, 2020.

7. Murthy S, Gomersall CD, Fowler RA. Care for Critically Ill Patients With COVID-19. JAMA. March 2020. doi:10.1001/jama.2020.3633

8. MacLaren G, Fisher D, Brodie D. Preparing for the Most Critically Ill Patients With COVID-19. JAMA. February 2020. doi:10.1001/jama.2020.2342

9. Kluge S, Janssens U, Welte T, Weber-Carstens S, Marx G, Karagiannidis C. Empfehlungen zur intensivmedizinischen Therapie von Patienten mit COVID-19. Medizinische Klin - Intensivmed und Notfallmedizin. March 2020:7–9. doi:10.1007/s00063-020-00674-3

10. Soar J et al. Adult advanced life support [12]. Resuscitation. 2015;95:100–147. doi:10.1016/j.resuscitation.2015.07.016

11. Ruan Q, Yang K, Wang W, Jiang L, Song J. Clinical predictors of mortality due to COVID-19 based on an analysis of data of 150 patients from Wuhan, China. Intensive Care Med. 2020. doi:10.1007/s00134-020-05991-x

